# Health Economic Analysis Plan for the Hospital Acquired Pneumonia PrEveNtion (HAPPEN) study: A multi-centre randomised control trial exploring the effectiveness of improving the frequency and quality of oral care in reducing the incidence of pneumonia

**DOI:** 10.1101/2025.08.25.25333724

**Authors:** Pakhi Sharma, Nicole M White, Nicholas Graves, Brett G Mitchell

## Abstract

The Hospital-acquired Pneumonia Prevention (HAPPEN) study is a multi-site, stepped wedge cluster randomised trial evaluating the impacts of improving oral care among adult patients to reduce the incidence of non-ventilator, hospital-acquired pneumonia (HAP). This document is the Health Economics Analysis Plan (HEAP) for evaluating the cost-effectiveness of the intervention compared with usual care.

The trial was preregistered on the Australian and New Zealand Clinical Trials registry (ACTRN12624000187549) and is funded by the Medical Research Future Fund (MRF2022645). A copy of the study protocol and a signed version of this plan are available on request from the corresponding author (BM)

## Trial title

Hospital Acquired Pneumonia PrEveNtion (The HAPPEN study): A multi-centre randomised control trial exploring the effectiveness of improving the frequency and quality of oral care, in reducing the incidence of pneumonia.

## Economic evaluation title

Economic evaluation of improved frequency and quality of oral care in reducing the incidence of hospital-associated pneumonia

## Trial registration number

The trial is registered with the Australia New Zealand Clinical Trial Registry (ANZCTR), registration number: <u>ACTRN12624000187549</u>

## Purpose of HEAP

The purpose of this HEAP is to describe the analysis and reporting procedure intended for the economic analyses to be undertaken for the HAPPEN study. The analysis plan is designed to ensure that there is no conflict with the protocol and associated statistical analysis plan and it should be read in conjunction with them.

## Trial protocol version

This document has been written based on information contained in the trial protocol version 1.3 dated 9 August 2024.

## Trial Statistical Analysis Plan (SAP) version

SAP Version: 1.3, Date: 7^th^ August 2025

## Trial HEAP version

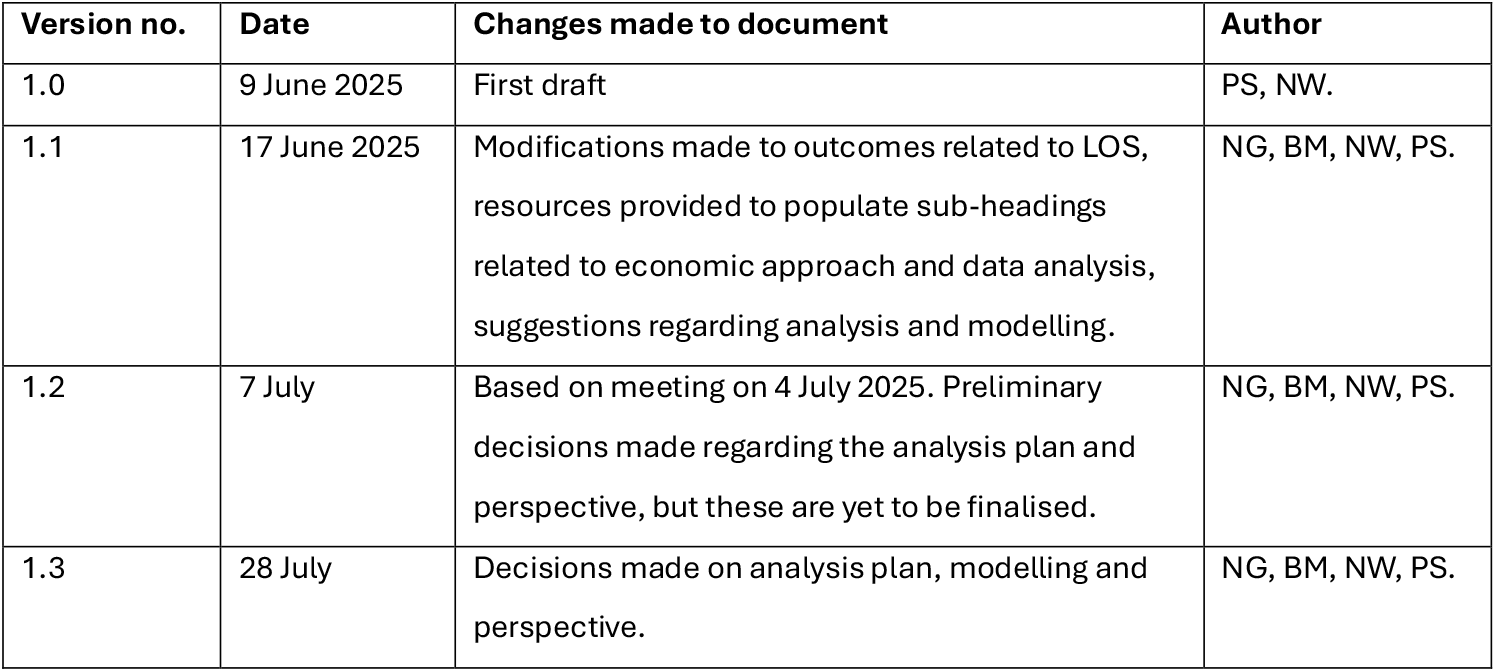

## Roles and responsibilities

The cost-effectiveness analysis was planned by the entire research team, led by PS, NW, BM and NG. The remaining CI and AI group to provide input, as needed. Meetings to progress the economic evaluation after the trial were planned and undertaken on an as-needed basis.

## Trial introduction and background

### Trial background and rationale

Hospital-associated pneumonia (HAP) is the most common hospital-acquired infections. They are a known issue in all hospital settings and contribute to increased lengths of stay, morbidity and mortality (1–3). Infection control, including quality and frequency of oral care, are a practical and important way to intervene in transmission, keeping patients safe (4). The Hospital Acquired Pneumonia PrEveNtion (HAPPEN) trial aims to explore the effectiveness and cost-effectiveness of this intervention. There is limited economic evidence available relating to the cost-effectiveness of improved frequency and quality of oral care in reducing the incidence of HAP in the Australian setting. A cost-effectiveness analysis will provide decision-makers with new and useful information that will help them make evidence-based funding allocation decisions.

A within-trial cost-effectiveness analysis was planned from the outset of the study and included in the approved trial protocol, where evaluating the intervention from an economic perspective was listed as a secondary outcome.

### Aim of the trial

Using an effectiveness-implementation hybrid design, the aim of this project is to prospectively evaluate the question “What is the effectiveness and cost-effectiveness of improving the frequency and quality of oral care, in reducing the incidence of HAP?”

### Objectives and/or research hypotheses of the trial

The **primary objective** of this study was to assess the clinical effectiveness of improving the frequency and quality of oral care, in reducing the incidence of HAP. One of the **secondary objectives** was to evaluate the cost-effectiveness of the HAPPEN intervention compared with usual care. Other objectives are listed in the trial protocol.

### Trial population

Three Australian hospitals categorised by the Australian Institute of Health and Welfare as either a principal referral hospital, public hospital (Group A) or private hospital (Group A) participated in the study. Three eligible wards were randomly selected, within each hospital.

Wards qualified for inclusion if they had one or more of the following characteristics enabling the recruitment of participants at greater risk of HAP: medical ward, stroke ward, patients with an average age greater than 70 years. Eligible wards must have a turnover of approximately 150 patients per month. Exclusions include any unit that cares for ventilated patients, paediatric, neonatal, adolescent, mental health, medical admission units and wards managing specific respiratory conditions e.g., COVID-19.

Patients (>18 years old) who are admitted to one of the wards on the day of the point prevalence survey will be used.

### Intervention(s) and comparator(s)

The intervention includes oral care, patient and staff education, and audit and feedback. The comparator is the current standard of care.

### Trial design

This was a multi-centre stepped-wedge cluster randomised trial. There were 9 clusters (three wards in each of the three hospitals) where a sequential rollout of the intervention was completed over 12 months. All wards received the intervention, with the timing randomised over three-monthly blocks. The shortest duration for the control arm is 3 months and the longest is 9 months. The study design allowed patients to benefit from the intervention.

### Trial start and end dates

Recruitment started on 3 June 2024. The trial was completed on 22 August 2025.

## Economic approach

### Aim(s) of economic evaluation

To determine whether a decision to adopt the Hospital Acquired Pneumonia PrEveNtion (HAPPEN) intervention is likely to be cost-effective as compared to usual care.

### Objective(s) of economic evaluation

The primary objective is to assess the cost-effectiveness of the HAPPEN intervention compared with usual care.

### Overview of economic analysis

The trial is conducted within the Australian healthcare context. Study setting is Australian hospital wards enrolled in the study. The within-trial economic evaluation will take place under the intention to treat principle. The analytical approach will take the form of cost-effectiveness analysis. A **hospital costing perspective** will be taken for the base-case analysis with all cost values reported in Australian dollars (year 2025). The time horizon will match the trial length i.e., 12 months. No discounting of costs or health outcomes will be undertaken if the time horizon does not exceed 12 months.

## Economic data collection & management

### Outcomes and measurement

The **primary health outcome** of this evaluation is adoption of the HAPPEN intervention. Changes to total costs and health benefits associated with adopting the intervention will be estimated using trial results and other hospital datasets. Changes to costs will include the resources required to implement the intervention and the changes to use of health services, for example, hospital bed days for treating HAP. Changes to health benefits will be measured using quality-adjusted life years (QALY) outcomes based on the estimated number of HAP cases prevented by data generated in the study (effectiveness).

### Data sources and collection

Existing literature and effectiveness outcomes derived from primary outcome of the trial will be used. The costs of adopting the intervention will be prospectively collected from hospital records. Excess attributable ward length of stay (LOS) that is associated with acquiring non-ventilator HAP will be sourced from the literature. There may be implementation costs on the following:

- **Staffing costs** (e.g., nursing) will be based on actual data collected from the trial, such as the number of nursing hours per day per ward. The cost estimates will include time allocated for staff education and support activities related to intervention delivery. Cost-effectiveness results will be presented per 1,000 hospital admissions.
- **Products cost**, e.g., consumables used in the intervention, including toothbrushes and toothpaste. Unit costs will be estimated per admission. The valuation of these resources will be based on market prices (5).

Data will be managed in accordance with the specifications outlined in the clinical trial protocol, and in compliance with all confidentiality, ethics, and governance requirements. Any data inputs required for the model that are not sourced directly from the trial will be identified and extracted following established evidence synthesis guidelines and relevant published literature. Expert opinion may be sought if existing literature is insufficient.

### Software

The decision analytic model will be programmed in TreeAge Pro Healthcare Version 2025 R1.1 and R version 4.5.1 or higher. The structure of the model will be decided upon by the entire research team which includes a health economist, the trial statistician, trial manager and numerous clinicians to ensure that it is a fair representation of reality.

## Economic data analysis

### Analysis population

Patients (>18 years old) who are admitted to one of the wards on the day of the point prevalence survey will be used. Patient inclusion and exclusion will be followed as per the plan outlined in the clinical trial protocol.

### Analysis plan

A within-trial analysis will be performed, taking a 12-month time horizon. Discounting will not be applied as the time horizon does not exceed 12 months.

Changes to health benefits will be estimated in QALYs using the number of life years gained from reduced infection outcomes, the mortality effects, and the valuation of the relevant health states obtained from the study. A **cost-effectiveness model** will be developed to include prior distributions on costs and effectiveness parameters. This will be evaluated using probabilistic sensitivity analysis to estimate the posterior distributions of ‘change to costs’ and ‘change to QALY’ outcomes. Modelled changes in the primary outcome (HAP) will be used to estimate changes in health benefits from cases prevented by the intervention. Change to costs associated with resource use, intervention costs and LOS will be compared between the intervention and the control groups. All parameters contributing to the cost outcomes for each option will be assigned uncertainty distributions. Monte Carlo sampling will be used to estimate the resulting cost outcomes (6).

The decision will be informed by plotting cost-effectiveness acceptability curves with threshold value between $0 and $100,000 per QALY gained. A maximum willingness to pay for marginal health benefits of $28,000 will be used based on recent Australian research (7). Cost-effectiveness will be summarised by the **incremental cost-effectiveness ratio (ICER)** and **net monetary benefit (NMB)**, which offer different summaries of the change in costs versus health benefits.

**Probabilistic sensitivity analysis** will be undertaken to account for uncertainty in model parameters and its impact on cost-effectiveness outcomes. Uncertainty in parameter estimates will be captured using appropriate statistical distributions to describe the variability (beta for transition probabilities; gamma or fixed for costs), the fitted distributions subject to random re-sampling via probabilistic sensitivity analysis.

For this model, only complete data will be used, with no missing data included. Data validation and cleaning will be undertaken, including face validity checks (e.g., identifying misspelled text) and verification against source documents. Any corrections made will be clearly documented. If substantial uncertainty remains around the intervention’s effectiveness, a value of information analysis will be conducted to assess whether further research would be worthwhile.

## Modelling

We will use a **cohort-level decision-analytic model** to estimate the outcomes of the trial. Cost-effectiveness will be shown using ICER, where the mean change to costs associated with the intervention is divided by the mean change in health outcomes, as well as NMB. Resource use, costs and health outcomes will be collected for all participants enrolled in the trial and will be used to parameterise the model. Secondary infection outcomes will be explored through scenario analyses. Subgroup analyses may be considered, depending on data availability and distribution, by hospital site, composite infection types, and/or per-protocol adherence.

The model structure maybe subject to change, following initial exploratory analysis of trial data. The model will be estimated and internally validated using trial data. The key assumptions of the model will be clearly defined. The incidence of HAP observed during the trial will be assumed to persist after the end of trial follow up.

## Reporting

The evaluation’s results will be reported according to the Consolidated Health Economic Evaluation Reporting Standards (CHEERS). Any deviation from HEAP will be described and justified in the final published report.

## Data Availability

All data produced in the present work are contained in the manuscript

## Abbreviations

Below is an alphabetical list of abbreviations present throughout this document.

ANZCTR: Australia New Zealand Clinical Trial Registry
CHEERS: Consolidated Health Economic Evaluation Reporting Standards
HAP: Hospital-acquired pneumonia
HAPPEN: Hospital Acquired Pneumonia PrEveNtion
ICER: Incremental cost-effectiveness ratio
LOS: Length of stay
MRFF: Medical Research Future Fund
NMB: Net monetary benefit
QALY: Quality adjusted life years
WTP: Willingness to pay

